# Perioperative Wearable Device Features are associated with Moderate-to-Severe Chronic Pain after Surgery in the “All of Us” Research Program

**DOI:** 10.1101/2025.09.23.25336489

**Authors:** Wenyu Zhang, Mindy K. Ross, Madelyn R. Frumkin

**Affiliations:** Department on Biomedical Data Science, Geisel School of Medicine at Dartmouth; Center for Technology and Behavioral Health, Dartmouth College; Departments of Psychiatry, Geisel School of Medicine at Dartmouth College

## Abstract

Chronic pain is a common and disabling condition affective over 50 million adults in the United States. Surgery may offer a critical opportunity to prevent chronic pain, as 10-35% of surgical patients develop new or worsening pain. However, prevention and treatment of chronic pain among surgical patients is hindered by a lack of reliable and clinically actionable biomarkers. Digital health technologies offer novel opportunities to develop and validate *digital biomarkers* of chronic pain among surgical patients. In this study, we leveraged data from the “All of Us” Research Program, which linked Electronic Health Record (EHR) data with participant’s own Fitbit data. We identified participants who: 1) had a surgical procedure as indicated in the EHR; 2) had Fitbit data available 30 days before and/or after surgery; and 3) completed the Overall Health Questionnaire assessing pain intensity 3 months to 5 years after surgery. Our final cohort included 302 surgical patients, 29% of whom reported moderate-to-severe pain approximately 1.5 years after surgery. Among the domain-specific models, sleep features provided the best predictive performance, achieving an AUC of .722 in a held-out test set. The lowest AIC was observed in the stepwise model based on the subset of participants whose Fitbits provided sleep stage data (n = 244, AIC = 178.75, AUC = .649, sensitivity = 0.37, specificity = 0.87). Younger age (OR = 0.97; p = 0.048), lower preoperative step variability (OR = 0.56; p = 0.009), and higher preoperative variability in REM sleep proportion (OR = 1.62; p = 0.023) were associated with greater risk of moderate-to-severe pain approximately 1.5 years after surgery. Digital biomarkers derived from consumer wearable device data appear highly promising for improving prediction and understand of chronic postoperative pain.

## 1. Introduction

Chronic pain (CP) is among the most common and disabling health conditions^1–3^. In the United States, over 50 million adults report pain on most days or every day^4^. Chronic pain conditions account for over $87 billion in annual healthcare spending, with costs increasing over the past several decades^5^. Individuals with chronic pain have increased risk of mental health problems, cognitive impairment, and worse overall health^1,6–8^.

Surgery is a common event that may offer key opportunities to prevent chronic pain. Over 300 million surgeries are performed worldwide each year^9,10^. Approximately 10-35% of surgical patients experience persistent post-surgical pain (PPSP), defined as pain that develops or increases in intensity after a surgical procedure and persists longer than 3 months^11–15^. PPSP is confined to the surgical area^15^. However, PPSP commonly co-occurs with other pain syndromes. Preoperative pain is among the strongest and most consistent risk factors for PPSP, especially preoperative pain outside the surgical area^16–18^. Surgical patients can also develop widespread chronic pain through changes to the central nervous system (CNS) that result in hypersensitivity to pain and other sensory stimuli, a phenomenon known as *central sensitization*^14,19,20^.

Prevention and treatment of CP among surgical patients is hindered by a lack of reliable and clinically-actionable biomarkers^21^. Although several models have been developed to identify patients at risk of PPSP, most have limited accuracy^16^. These models rely primarily on demographic and patient-reported information, including pre-existing pain and psychosocial symptoms. Digital health technologies (DHTs) offer novel opportunities to develop and validate *digital biomarkers* of chronic pain among surgical patients. The Food and Drug Administration (FDA) defines a digital biomarker as, “a characteristic or set of characteristics, collected from digital health technologies, that is measured as an indicator of normal biological processes, pathogenic processes, or responses to an exposure or intervention, including therapeutic interventions.”^22^ Digital biomarkers derived from smartphones and wearable devices have proven highly valuable for predicting key health outcomes, including obesity, sleep apnea, major depressive disorder, and all-cause mortality^23–25^. In the surgical domain, wearables have primarily been applied to monitoring postoperative recovery via physical activity^26,27^. There is significant untapped potential to measure and monitor hypothesized mechanisms of chronic widespread pain, including both pre- and postoperative physical activity, sleep, and cardiovascular functioning^28–34^. Importantly, self-report of these constructs tends to have low agreement with objective measurement^35–38^. DHTs can therefore increase precision of model inputs via continuous objective assessment of factors hypothesized to play a key role in CP.

The National Institutes of Health’s “All of Us” Research Program (AoURP) is a large, longitudinal cohort that integrates Fitbit data with electronic health records (EHRs) and patient-reported outcomes^39^. Studies leveraging AoURP have examined associations between wearable-derived activity and sleep metrics and a range of outcomes, including chronic disease, mental health conditions, and surgical recovery^10–12^. In the surgical domain, recent investigations using the “All of Us” data suggest utility of wearable device data for identifying patients at risk of postoperative complications, acute postoperative pain, and chronic opioid use^42,43^. Despite these advances, little is known about whether perioperative digital biomarkers can identify individuals at risk for moderate-to-severe chronic pain after surgery. Thus, the goals of this study were to: (i) derive peri-operative Fitbit features across multiple domains, (ii) evaluate their associations with moderate-to-severe chronic pain after surgery, and (iii) assess the predictive performance of digital biomarkers in independent test data.

## 2. Methods

### 2.1 Study Design and Cohort Selection

This retrospective observational study drew on the Controlled Tier dataset (version 8) of the AoURP. Eligible participants had a surgical procedure recorded in their EHR, contributed at least five valid days of Fitbit data within 61 days before or after that procedure, and completed the Overall Health Survey between 3 months (≥ 90 days) and 5 years (<1,800 days) after surgery. When participants underwent multiple operations, we selected the most recent procedure meeting these criteria. Participants who reported a past week average pain intensity ≥ 4 out of 10 in the Overall Health Survey were categorized as having moderate-to-severe pain.

### 2.2 Feature Derivation

Wearable features were extracted from daily Fitbit summaries in three domains: physical activity, resting heart rate, and sleep. Features were separately computed for the pre- and postoperative periods. Participants needed at least five valid days of data in each domain to contribute to following analyses. A valid day was defined as a day with recorded, non-missing values for total step count (> 0), average resting heart rate, and total sleep duration. For each domain and period, we computed three statistics: the mean value across days, the standard deviation (SD) across days, and a linear slope over time obtained by fitting a simple regression to the daily measurements within the 30-day window. Continuous variables were scaled to the 0-1 range using min-max normalization to facilitate comparisons across measurement units.

Physical activity features included the mean number of steps per day, the SD of daily steps and the slope of steps over time. The number of valid days was recorded to reflect data completeness. Resting heart rate features comprised the mean daily value, the SD of daily values and the slope over time, together with the number of valid days. Sleep was recorded in either Classic or Stages mode. For both modes we derived total sleep duration (minutes asleep excluding wakefulness), sustained awakenings (continuous time awake) and restlessness (brief arousals below the wake threshold). Sleep efficiency, defined as the ratio of total sleep duration to time in bed, and sleep fragmentation, defined as the proportion of time in bed spent awake or restless, were calculated for each night and summarized across the pre- and postoperative periods by their mean, SD and slope. Among participants with Stages-mode data, additional variables described the distribution of sleep across physiologic states; Fitbit estimates the minutes spent in rapid eye movement (REM), light and deep sleep. The proportion of total sleep time spent in each stage was calculated, and its mean, SD and slope were derived. To minimize the influence of extreme sleep values, individuals whose total sleep duration fell outside the interquartile range for either the preoperative or postoperative period were excluded. The detailed feature list is shown in **Supplemental Table 1**.

### 2.3 Covariates and Missing Data

Age, sex, and race were included as demographic covariates. Race was categorized to a binary variable (White vs. non-White) due to small cell counts among non-White racial groups. Under the All of Us Data and Statistics Dissemination Policy, cells that correspond to fewer than 20 participants must be collapsed or aggregated. The response time between surgery and survey completion, measured in months, was used to adjust for differences in follow-up time.

Demographic variables were complete, whereas a small proportion of domain-specific Fitbit features were missing; these values were imputed using multivariate imputation by chained equations with predictive mean matching, as implemented in the mice() package in R.

### 2.4 Predictive Modeling and Evaluation

The analytic workflow is outlined in **Figure 1**. In the first step, each wearable-derived feature was examined in a univariate logistic regression adjusted for age, sex, race and survey response time, with CP after surgery as the outcome. Features were assessed separately for preoperative and postoperative periods, and those meeting a significance threshold of p < 0.05 were flagged for further consideration.

**Figure 1.**
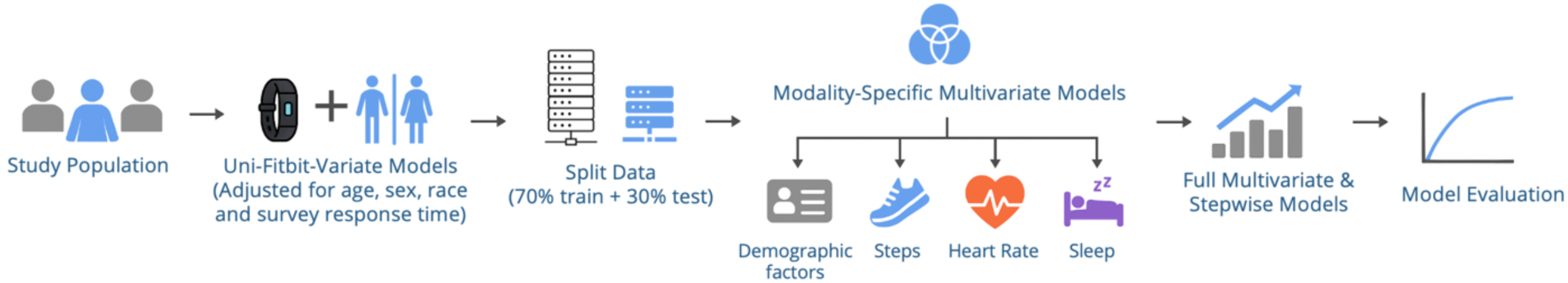
Overview of the modeling workflow for postoperative chronic pain prediction using perioperative Fitbit, EHR and survey data.

We then built domain-specific multivariable models, including: 1) a demographics-only model (baseline); 2) a model incorporating physical activity features; 3) a model incorporating heart rate features; 4) a model incorporating sleep features, based on all available data; and 5) a model incorporating sleep features, based on the subset of participants who Fitbit provided information on sleep stages. Each model adjusted for the demographic covariates and included all candidate features from the relevant domain. These models assessed whether individual features remained independently associated with CP when analyzed jointly.

To explore the added value of combining modalities, significant features from the univariate analyses across all domains were pooled into comprehensive multivariable logistic regression models. Model selection was guided by the Akaike Information Criterion (AIC) using a stepwise procedure to produce parsimonious models while limiting multicollinearity. The data set was randomly divided into a training set (70%) and a hold-out test set (30%). Predictive performance was quantified using the area under the receiver operating characteristic (ROC) curve (AUC) on the test data.

To quantify feature importance and enhance interpretability, we systematically perturbed individual features and computed the change in predicted probability from the original prediction. This delta probability (Δ-prob) reflects the marginal impact of each feature on the model output^44^. Values were aggregated across observations and summarized as the mean absolute contribution per feature. Features were ranked by mean absolute delta probability, and the top 5 most influential features were shown for visualization. These filtered features were displayed in jittered scatter plots showing the distribution of Δ-prob values for each feature, with annotations of average feature importance scores.

## 3. Results

Among the 1,688 individuals with a valid surgery record and Fitbit coverage, 757 completed the Overall Health Survey within 90 days to five years after surgery. After applying the requirement of at least five valid days of step, resting heart rate, and sleep data, the final analytic cohort included 302 participants, of whom 87 (28.8%) were classified as moderate-to-severe pain group and 215 (71.2%) as no-to-low pain group. A subset of 244 participants had stage-mode devices that provided detailed sleep stage data (**Figure 2**).

**Figure 2.**
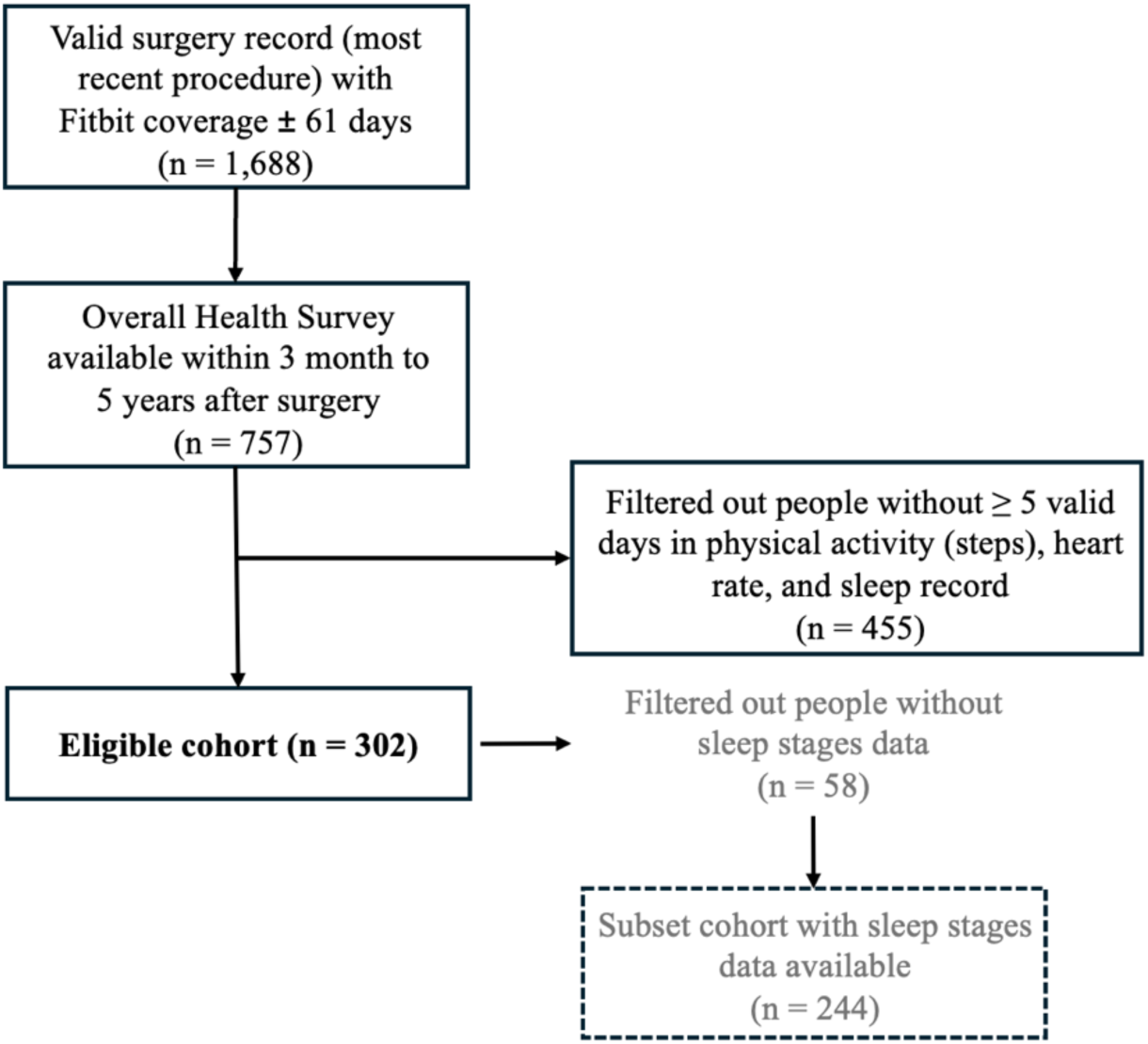
Cohort Selection Flowchart.

Participants with moderate-to-severe pain after surgery were younger on average than those with no-to-low pain (53.36 vs. 57.12 years; p = 0.032) and more likely to identify as non-White (21.84% vs. 12.09%; p = 0.048). A higher proportion of women were observed in the moderate-to-severe pain group (79.31% vs. 67.90%), although this difference was not statistically significant (p = 0.066). Survey response time was comparable between groups (17.26 vs. 18.16 months; p = 0.615).

Distributions of pre- and postoperative values stratified by pain group are shown in **Figure 3**, with descriptive statistics presented in **Supplemental Table 2**. Univariate logistic regression models adjusted for demographics and survey response time identified several Fitbit-derived features that were significantly associated with moderate-to-severe postoperative pain (**Table 1**). Lower mean step counts and reduced step variability, both before and after surgery, were associated with higher odds of moderate-to-severe pain. Elevated postoperative resting heart rate and greater variability in resting heart rate also predicted increased risk. For sleep, shorter total duration and greater postoperative variability in duration, efficiency, and fragmentation were significant predictors. Among participants with stage-mode devices, lower REM sleep proportion and greater variability in REM (pre- and post-operatively), light (preoperatively), and deep (postoperatively) sleep were also associated with higher odds of moderate-to-severe pain.

**Figure 3.**
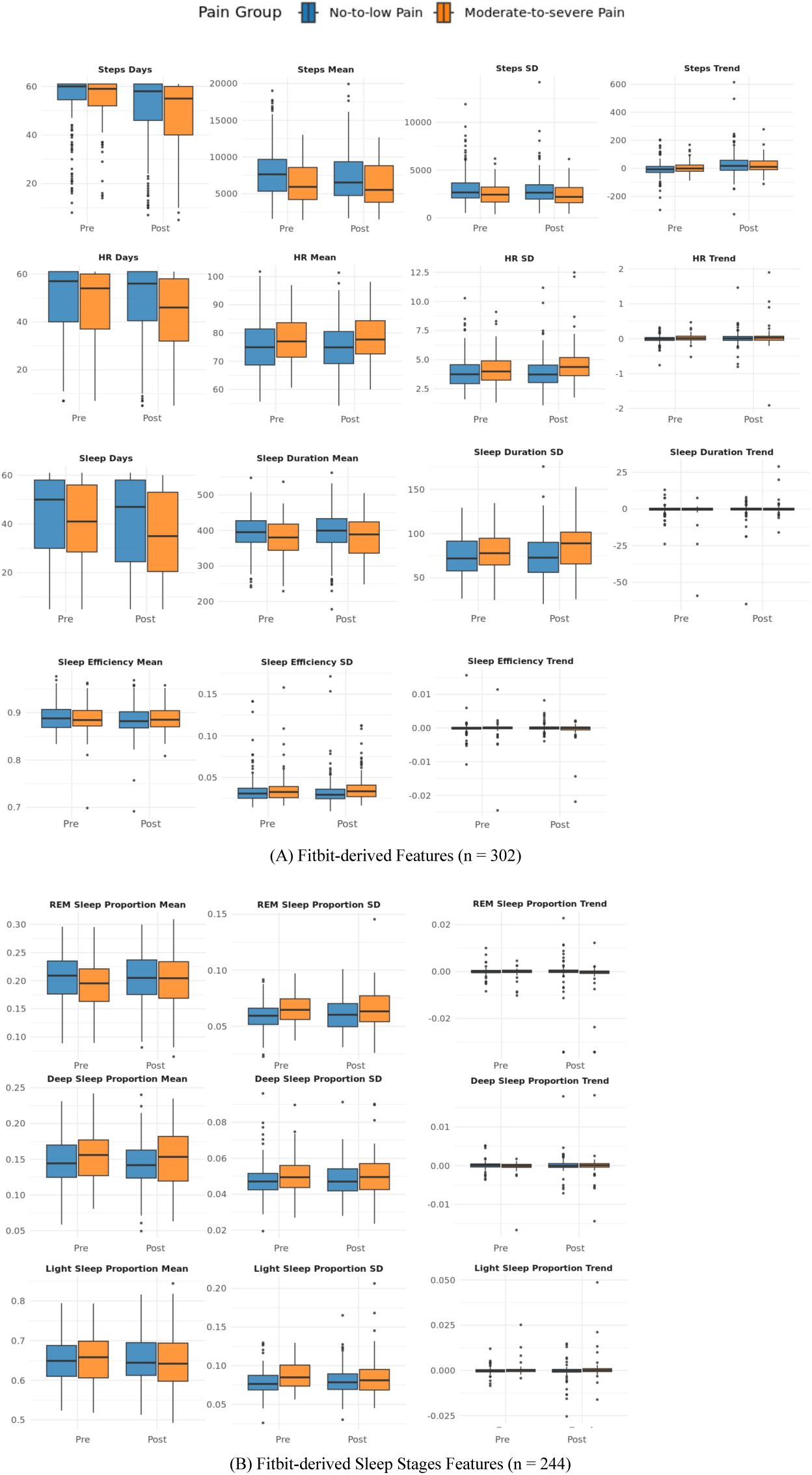
Boxplots of Fitbit-derived features (mean and SD) in no-to-low pain and moderate-to-severe pain groups across pre- and postoperative periods. (A) displays physical activity, resting heart rate, and sleep features for the full analytic cohort (n = 302). (B) shows sleep stage-specific features among participants with stage-mode devices (n = 244). Units: Steps = daily steps; HR = Resting Heart Rate (beats per minute); Sleep Duration = hours; Sleep Efficiency = proportion; Sleep Fragmentation = proportion; REM/Deep/Light Sleep = proportion.

**Table 1.** Univariate logistic regression results for predicting persistent chronic pain, adjusted for demographics and survey response time, displaying variables with statistical significance.

| Modality | Variable | ORs | 95% CI | p-value |
| --- | --- | --- | --- | --- |
| **Physical Activity** | Steps Mean (pre) | 0.63 | 0.47-0.85 | 0.002 |
|  | Steps SD (pre) | 0.67 | 0.49-0.91 | 0.012 |
|  | Steps Mean (post) | 0.71 | 0.53-0.94 | 0.017 |
|  | Steps SD (post) | 0.68 | 0.50-0.94 | 0.020 |
| **Resting Heart Rate** | Mean (post) | 1.38 | 1.05-1.82 | 0.023 |
|  | SD (post) | 1.61 | 1.23-2.11 | <0.001 |
| **Sleep** | Total Duration Mean (pre) | 0.71 | 0.54-0.92 | 0.010 |
|  | Total Duration Mean (post) | 0.71 | 0.54-0.92 | 0.010 |
|  | Total Duration SD (post) | 1.53 | 1.17-2.00 | 0.002 |
|  | Efficiency SD (post) | 1.53 | 1.16-2.02 | 0.018 |
|  | Fragmentation SD (post) | 1.37 | 1.05-1.79 | 0.038 |
|  | Days Available (post) | 0.76 | 0.59-0.98 | 0.018 |
| **Sleep Stages**<br>**(n = 244)** | REM Sleep Mean (pre) | 0.66 | 0.49-0.89 | 0.007 |
|  | REM Sleep SD (pre) | 1.96 | 1.43-2.70 | <0.001 |
|  | REM Sleep SD (post) | 1.49 | 1.11-2.00 | 0.008 |
|  | Light Sleep SD (pre) | 1.76 | 1.30-2.39 | <0.001 |
|  | Deep Sleep SD (post) | 1.41 | 1.07-1.86 | 0.015 |

Physical activity, heart rate, and sleep metrics are based on the full cohort ( $n = 302$ ), while sleep stage features are limited to participants with available stage data ( $n = 244$ ). Each feature was scaled to the 0-1 range using min-max normalization and evaluated by univariate logistic regression model, adjusted for age, sex, race, and survey response time. SD denotes standard deviation and trend reflects the linear slope over time. The odds ratios (ORs), 95% confidence intervals (CIs), and p-values are reported.

**Figure 4** shows ROC curves and feature importance for domain-specific models. The demographic-only baseline model had a test AUC of .584 with poor sensitivity (.032). Step count features improved performance (test AUC = .677, sensitivity = .10, specificity = .95), with preoperative mean steps, postoperative step trend, and step data availability contributing most strongly. Resting heart rate features produced a test AUC of .596 (sensitivity = .23, specificity = .90), with postoperative mean resting heart rate, variability in resting heart rate, and preoperative mean heart rate as the greater importance. Among domain-specific models, classic sleep features achieved the highest discrimination (test AUC = .722, sensitivity = .42, specificity = .92), with preoperative sleep efficiency, postoperative sleep fragmentation, and variability in sleep duration and fragmentation as the most important predictors. Sleep stage features yielded a test AUC of .652 (sensitivity = .19, specificity = .87), with REM and light sleep proportions in both pre- and postoperative periods as the dominant predictors. Performance metrics for all models are summarized in **Supplemental Table 3**.

**Figure 4.**
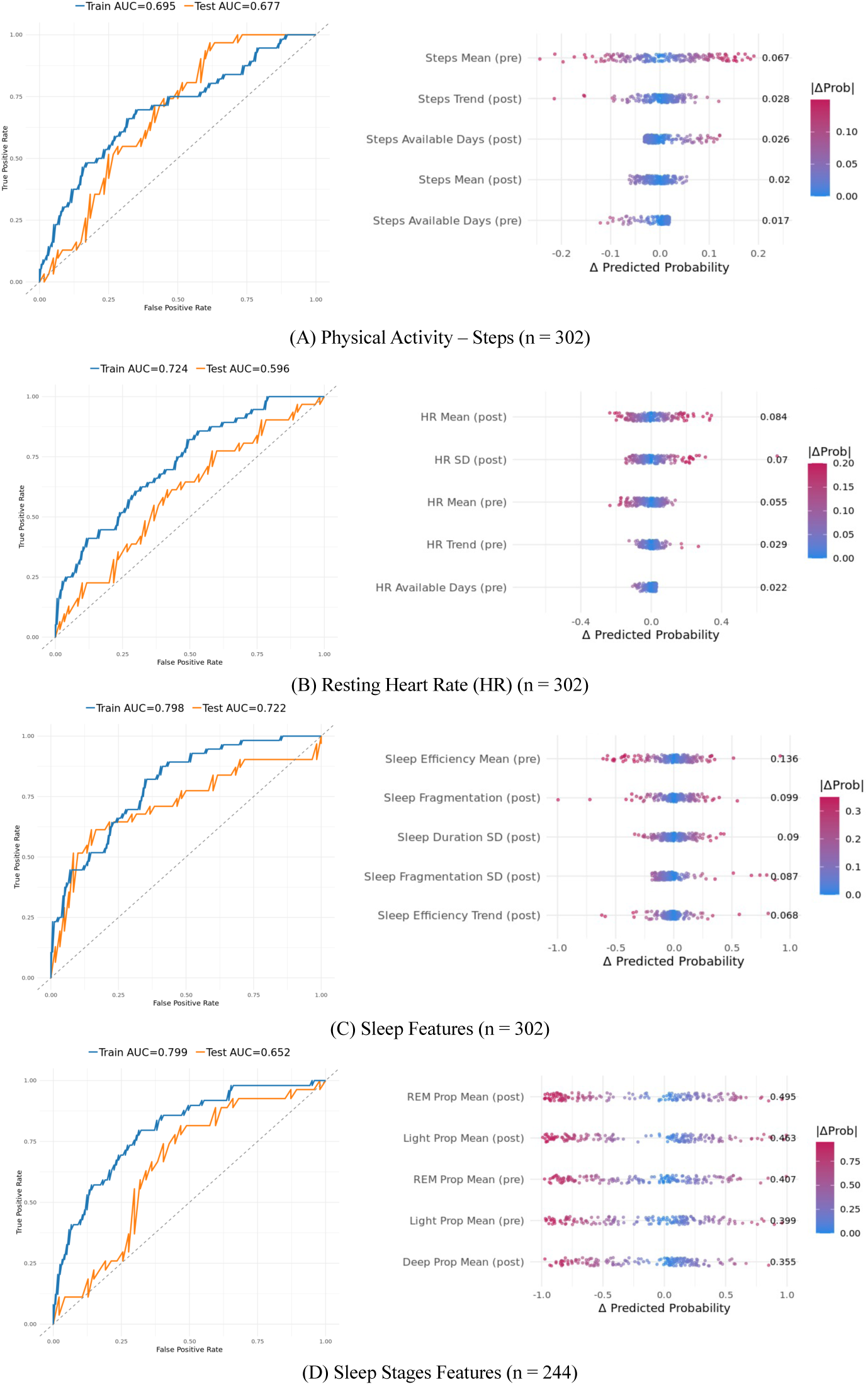
Modality-specific model ROC curves and feature importance. Four logistic regression models were trained to predict chronic pain using single-modality features: (A) physical activity, (B) resting heart rate, (C) classic sleep features, and (D) sleep stage features. Left panels show ROC curves for training (blue) and testing (orange) sets with corresponding AUCs. Right panels show SHAP-like feature importance plots, where each point represents a participant and color indicates the magnitude of the predicted probability change (|ΔProb|) when the feature is neutralized; features are ordered by mean absolute impact.

In the full cohort (n = 302), the multivariate regression achieved a test AUC of .679 (sensitivity = 0.26, specificity = 0.92), with preoperative and postoperative step means, postoperative variability in resting heart rate, survey response time, and postoperative step variability emerging as the most influential predictors. In the stage-mode cohort (n = 244), the model yielded a test AUC of .671 (sensitivity = 0.44, specificity = 0.87), with key contributors including pre- and postoperative step means, age, preoperative REM sleep proportion variability, and postoperative variability in resting heart rate (**Figure 5**).

**Figure 5.**
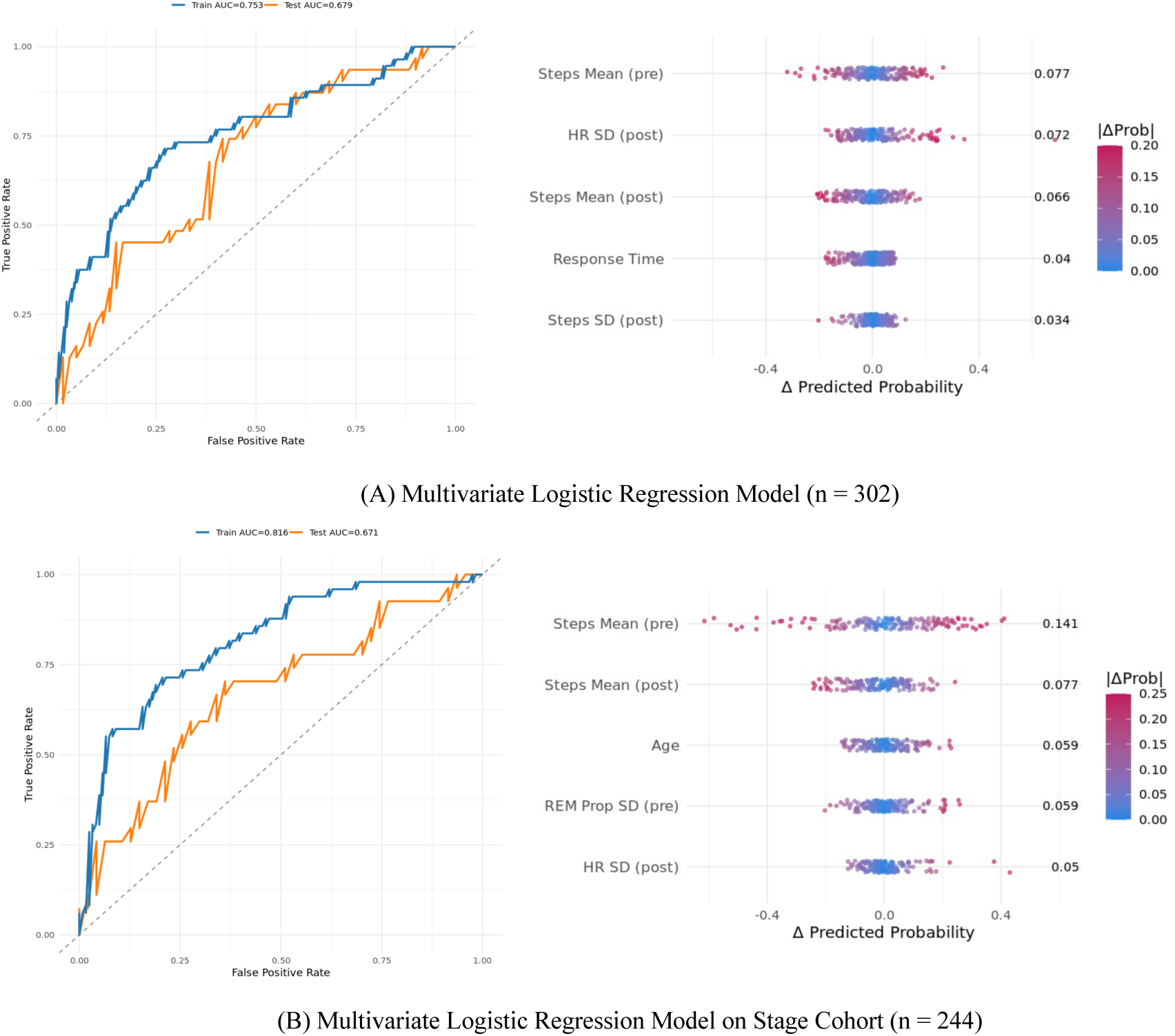
Multivariate logistic regression model ROC and SHAP-like curves for (A) full cohort (n = 302) and (B) participants with stage-mode data (n = 244).

Stepwise reduction lowered test AUCs modestly (0.648 and 0.649, respectively) while improving model parsimony. The lowest AIC was observed in the stepwise model based on the subset of participants whose Fitbits provided sleep stage data (AIC = 178.75, AUC = .649, sensitivity = 0.37, specificity = 0.87). Stepwise feature selection identified independent predictors retained in the final multivariate logistic regression models (Table 3). In the stage- mode cohort (n = 244), significant risk predictors were younger age (OR = 0.97; p = 0.048), lower preoperative step variability (OR = 0.56; p = 0.009), and higher preoperative variability in REM sleep proportion (OR = 1.62; p = 0.023). In the full cohort (n = 302), significant predictors included lower preoperative step variability (OR = 0.62; p = 0.020) and greater postoperative variability in resting heart rate (OR = 1.87; p = 0.004).

## 4. Discussion

Recent surveys suggest 30-44% of adults in the US own wearable devices^45,46^. These devices are low-cost and highly scalable sources of health information. Despite increasing use of wearable devices to monitor postoperative recovery^26,47^, few studies have leveraged preoperative wearable device data to identify novel digital biomarkers associated with postoperative outcomes, such as chronic postoperative pain. In this study, we leveraged publicly available data from the “All of Us” Research Program (AoURP) to identify perioperative digital biomarkers associated with moderate-to-severe chronic pain after surgery. Among the domain-specific models, we observed that sleep features provided the best predictive performance, achieving and AUC of .722 in a held-out test set. Notably, this model derived solely from demographics and consumer wearable device data performed as well as some existing prospective models of PPSP, which rely on patient-reported inputs^16^.

In examining feature importances through perturbation, preoperative sleep efficiency contributed the most to model accuracy in the domain-specific model. Higher preoperative variability in REM sleep proportion was also associated with greater likelihood of moderate-to-severe chronic pain after surgery among participants whose Fitbit devices provided sleep stage data. This feature remained among the most influential in the multivariate model, which was selected as the best-performing model based on the AIC, which balances model complexity with predictive performance.

Chronic pain conditions are frequently associated with sleep disturbance^48^. Given that we were not able to assess preoperative chronic pain in this retrospective analysis, it is possible our findings can be explained by worse sleep among individuals with pre-existing pain, who are at greater risk of persistent chronic pain after surgery^13^. However, mounting evidence suggests impaired sleep can cause or modulate acute and chronic pain^48–51^. In the surgical domain, there are highly mixed findings regarding a potential association between preoperative sleep disturbances and PPSP^37^. Importantly, existing studies primarily rely on patient-report of sleep characteristics, which have been shown to have very low agreement with objective measurement^36^. Our findings highlight opportunities for leveraging widely available consumer wearables to understand the role of sleep in PPSP.

Although domain-specific models based on sleep data out-performed models based on physical activity and heart rate data, features derived from these domains were highlighted in the stepwise models. Preoperative variability in average daily step count emerged as a consistent significant predictor between both cohort models (participants with and without sleep stage data). Prior research using the AoURP data set suggests low preoperative step counts (e.g., <7,500 steps per day) are associated with increased risk of postoperative complications^43^. Lower levels of physical activity are also associated with increased risk of developing neck, low back, and hip pain^52^. In univariate analyses, we similarly observed lower average preoperative daily step counts were associated with increased risk of moderate-to-severe chronic pain after surgery. In multivariate stepwise models, day-to-day *variability* in these values emerged as more influential. Wearable device data are longitudinal, offering novel opportunities to investigate symptom dynamics in addition to person-level averages. Our findings suggest such dynamic may have meaningful implications. For example, perhaps greater day-to-day variability in physical activity suggests engagement in physical therapy or attending work. Future research can leverage location data to better contextualize activity patterns.

We similarly observed that greater day-to-day variability in resting heart rate across the postoperative period was associated with increased risk of moderate-to-severe chronic pain after surgery. Resting heart rate is broadly understood as a key marker of cardiovascular functioning and metabolic demand^53^. Surgery and the subsequent acute recovery process involve a cascade of physiological responses that increase the body’s energy requirements. In the current study, greater variability in postoperative resting heart rate likely reflects greater impact of surgery on cardiac autonomic function. This could be due to more involved surgical procedures, which increase risk for PPSP^14^. However, it is also notable that autonomic functioning has been shown to vary across individuals, with implications for response to acute pain^54^. Thus, remote postoperative heart rate monitoring via consumer wearable devices may enhance early identification of patients at risk for chronic pain after surgery.

Importantly, this study has several limitations. Based on how pain was assessed in the AoURP, we were unable to determine whether pain reported 3+ months after surgery was related to the surgery itself (i.e., PPSP). Consequently, we do not characterize our findings as offering definitive insights into PPSP, which is defined as being localized to the surgery site^15^. Similarly, we did not have pre-surgery pain ratings available in this retrospective data set. Given that preoperative pain is a strong risk factor of postoperative pain^13,16^, our study may have identified correlates of chronic pain generally. Furthermore, surgery type was very heterogeneous in our sample and in some cases was hard to categorize due to limitations of available EHR data. Many individuals were also filtered out of the analysis due to incompatible timing of the Overall Health Questionnaire, resulting in a substantial loss of data. The sample size was relatively small for predictive modeling, and the study population was predominantly White. It has been previously reported that 80% of participants who provide their own Fitbit data in the AoURP identify as White, and 88% identifying as Non-Hispanic or Latino^55^. Going forward, AoURP plans to address this gap by providing participants with wearable devices, rather than relying on participants’ own device data. Given that Blacks/African American adults tend to experience more severe postoperative pain^56^, further research in larger, more diverse samples is critical.

Our analysis focused on daily summary measures of steps, resting heart rate, and sleep. More granular time-series data (e.g., hourly or intra-night patterns) were not explored due to challenges with accessing this data in the AoURP platform but may provide additional insights. Given rapid advancements in consumer wearable devices over the past few years, it is also notable that some features of interest are not yet available in the current data. For example, we highlight our findings related to day-to-day variability in resting heart rate. Modern consumer wearables can calculate heart rate variability (HRV), which indexes the variation in time between consecutive heartbeats. HRV is hypothesized to be highly relevant to acute and chronic pain conditions, but has thus far been primarily studied in highly-controlled laboratory settings^57,58^. New generation consumer wearables will offer key opportunities to evaluate a wider array of physiological features as predictors of PPSP.

Finally, although our models tended to have high specificity, sensitivity was low. In terms of resource allocation, it is beneficial to have a low number of false positives (high specificity). Lower sensitivity was somewhat expected given that less than half the sample exhibited moderate-to-severe chronic pain. Indeed, prior prediction models of PPSP have found a similar pattern of findings^59^. However, it will be important to address this gap in future research.

Future research should validate our findings in larger, more diverse, and well-characterized cohorts of surgical patients. Primarily, prospective studies are needed to determine whether perioperative digital biomarkers have clinical utility for the prognostic prediction of PPSP. These studies should adhere to guidelines from the IASP to define PPSP as new or worsening pain at the surgical site, although prediction of new or worsening widespread chronic pain is also of interest given the significant functional and psychosocial impacts of widespread pain conditions. Access to raw wearable device data will facilitate investigation of novel features (e.g., HRV) and may improve accuracy. Access to raw data will also facilitate the development of open-source, reproducible pipelines for cleaning and analyzing complex wearable device data. Additionally, prior research suggests integration of objective wearable device data with patient-reported factors (e.g., psychosocial symptoms) provides greater predictive accuracy^60^. In future studies with larger samples, modern artificial intelligence/machine learning techniques including gradient boosting machines, random forests, deep neural networks, and ensemble models may also improve model performance.

## 5. Conclusion

Overall, our findings suggest that consumer wearables provide a scalable approach to improve prediction and understanding of chronic pain after surgery. Integration of wearable device data with clinical and patient-reported information may improve risk stratification and enable earlier identification of patients at risk of PPSP. Wearable devices can also facilitate greater understanding of biopsychosocial mechanisms of acute-to-chronic pain transitions after surgery. Validation of these techniques in larger and more diverse prospective cohorts will be important to advance risk prediction and guide interventions for PPSP.

## Data Availability

All data produced in the present study are available through the All of Us Research Hub.

## Acknowledgments

The All of Us Research Program is supported by the National Institutes of Health, Office of the Director: Regional Medical Centers: 1 OT2 OD026549; 1 OT2 OD026554; 1 OT2 OD026557; 1 OT2 OD026556; 1 OT2 OD026550; 1 OT2 OD 026552; 1 OT2 OD026553; 1 OT2 OD026548; 1 OT2 OD026551; 1 OT2 OD026555; IAA #: AOD 16037; Federally Qualified Health Centers: HHSN 263201600085U; Data and Research Center: 5 U2C OD023196; Biobank: 1 U24 OD023121; The Participant Center: U24 OD023176; Participant Technology Systems Center: 1 U24 OD023163; Communications and Engagement: 3 OT2 OD023205; 3 OT2 OD023206; and Community Partners: 1 OT2 OD025277; 3 OT2 OD025315; 1 OT2 OD025337; 1 OT2 OD025276. In addition, the All of Us Research Program would not be possible without the partnership of its participants.

---

Appendix

**Supplemental Table 1.** Overview of Fitbit-derived features, including category, feature definition, metrics, and data availability.

| Category | Feature | Metric | Description | Availability |
| --- | --- | --- | --- | --- |
| **Physical Activity** | Step count | Mean, SD, Slope | Total steps per day | All |
| **Physiology** | Resting heart rate | Mean, SD, Slope | Average daily heart rate | All |
| **Sleep** | duration | Mean, SD, Slope | Minutes asleep per day | All |
|  | Efficiency | Mean, SD, Slope | Proportion of time asleep while in bed | All |
|  | Fragmentation | Mean, SD, Slope | % time awake/restless/wake while in bed | All |
|  | REM stage proportion | Mean, SD, Slope | Proportion of sleep spent in REM stage | Stages only |
|  | deep stage proportion | Mean, SD, Slope | Proportion of sleep spent in deep stage | Stages only |
|  | light stage proportion | Mean, SD, Slope | Proportion of sleep spent in light stage | Stages only |

**Supplemental Table 2.**
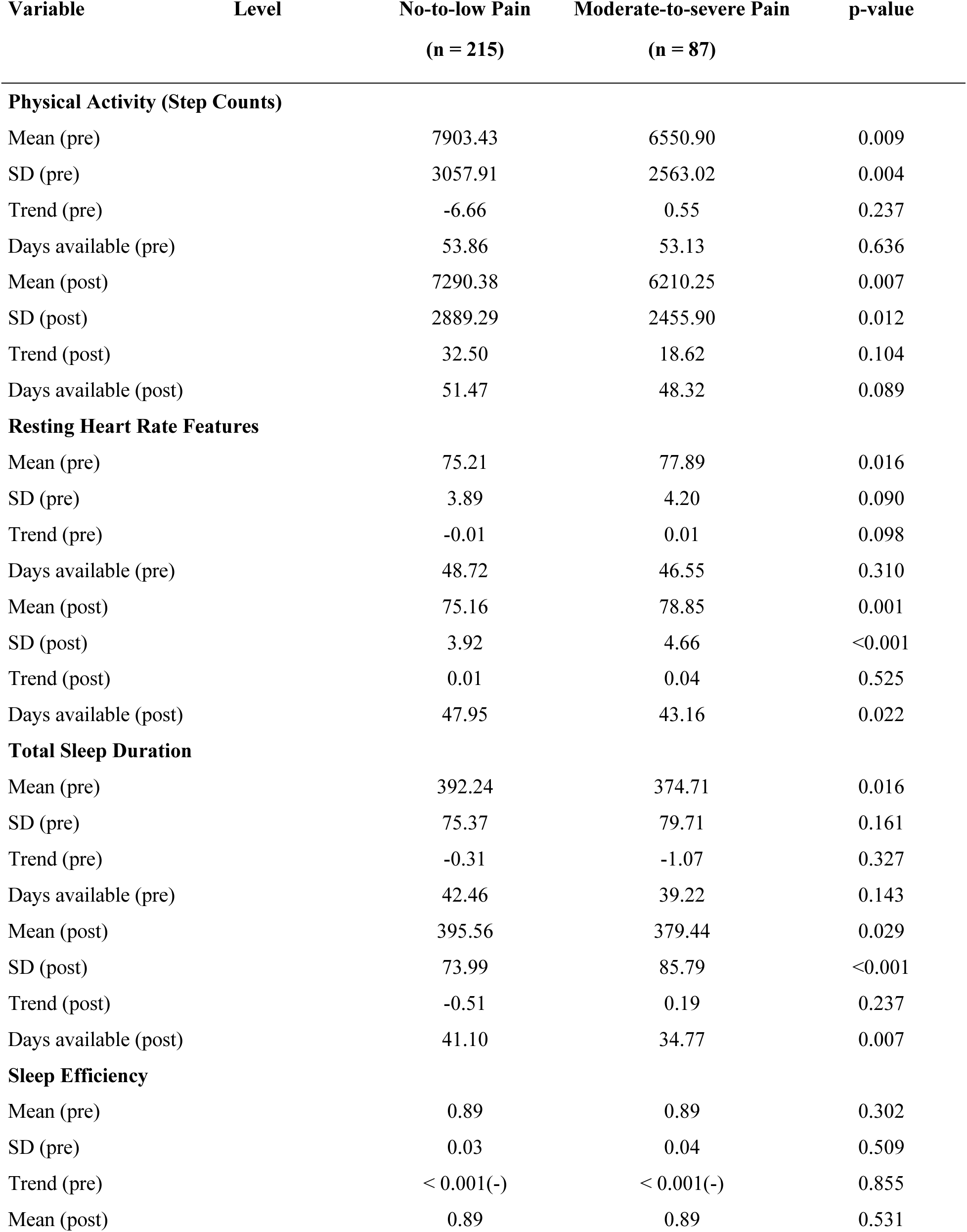
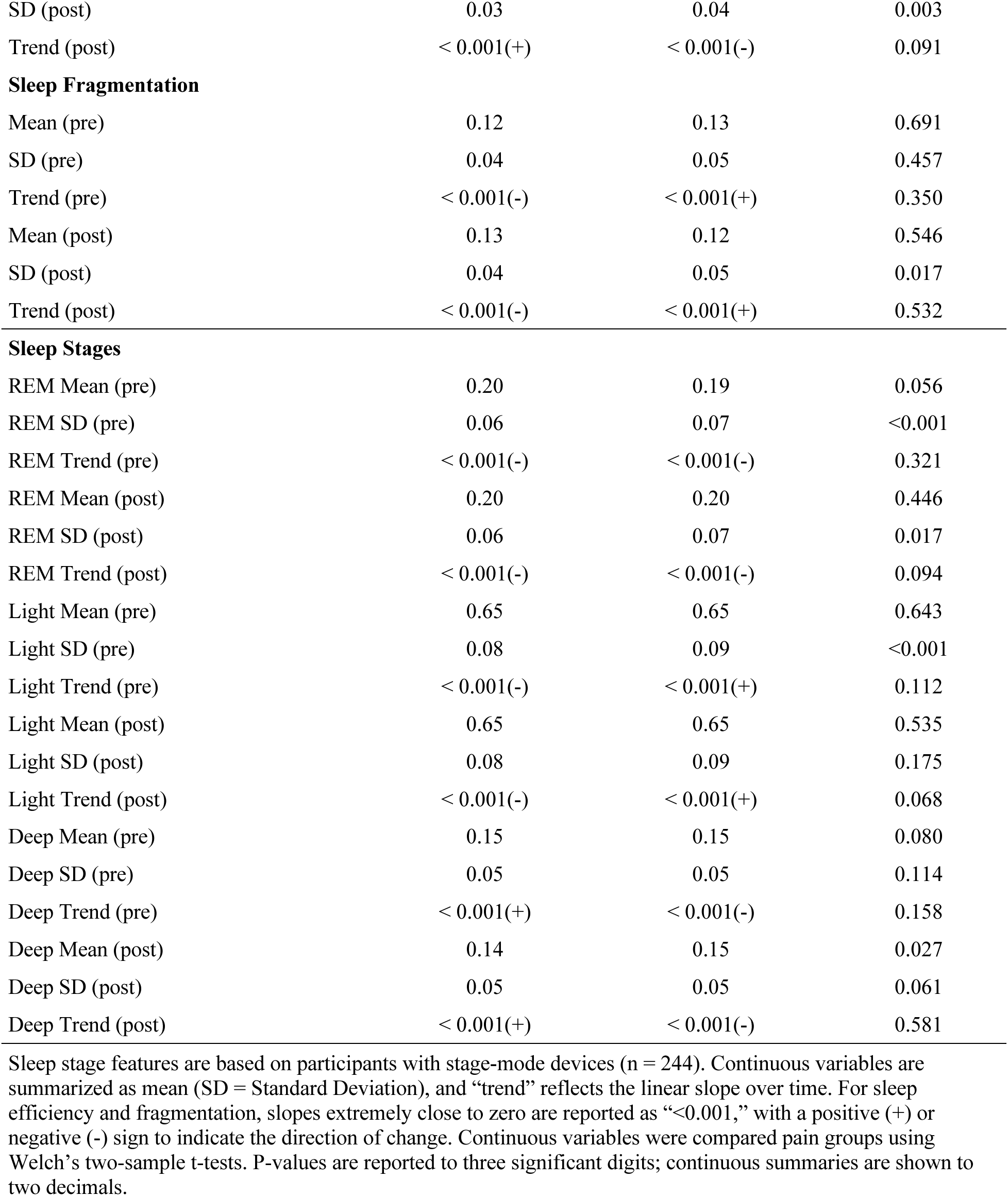
Descriptive statistics of Fitbit-derived features in the pre- and postoperative periods, stratified by pain group.

| Variable | Level | No-to-low Pain<br>(n = 215) | Moderate-to-severe Pain<br>(n = 87) | p-value |
| --- | --- | --- | --- | --- |
| **Physical Activity (Step Counts)** |  |  |  |  |
| Mean (pre) |  | 7903.43 | 6550.90 | 0.009 |
| SD (pre) |  | 3057.91 | 2563.02 | 0.004 |
| Trend (pre) |  | -6.66 | 0.55 | 0.237 |
| Days available (pre) |  | 53.86 | 53.13 | 0.636 |
| Mean (post) |  | 7290.38 | 6210.25 | 0.007 |
| SD (post) |  | 2889.29 | 2455.90 | 0.012 |
| Trend (post) |  | 32.50 | 18.62 | 0.104 |
| Days available (post) |  | 51.47 | 48.32 | 0.089 |
| **Resting Heart Rate Features** |  |  |  |  |
| Mean (pre) |  | 75.21 | 77.89 | 0.016 |
| SD (pre) |  | 3.89 | 4.20 | 0.090 |
| Trend (pre) |  | -0.01 | 0.01 | 0.098 |
| Days available (pre) |  | 48.72 | 46.55 | 0.310 |
| Mean (post) |  | 75.16 | 78.85 | 0.001 |
| SD (post) |  | 3.92 | 4.66 | <0.001 |
| Trend (post) |  | 0.01 | 0.04 | 0.525 |
| Days available (post) |  | 47.95 | 43.16 | 0.022 |
| **Total Sleep Duration** |  |  |  |  |
| Mean (pre) |  | 392.24 | 374.71 | 0.016 |
| SD (pre) |  | 75.37 | 79.71 | 0.161 |
| Trend (pre) |  | -0.31 | -1.07 | 0.327 |
| Days available (pre) |  | 42.46 | 39.22 | 0.143 |
| Mean (post) |  | 395.56 | 379.44 | 0.029 |
| SD (post) |  | 73.99 | 85.79 | <0.001 |
| Trend (post) |  | -0.51 | 0.19 | 0.237 |
| Days available (post) |  | 41.10 | 34.77 | 0.007 |
| **Sleep Efficiency** |  |  |  |  |
| Mean (pre) |  | 0.89 | 0.89 | 0.302 |
| SD (pre) |  | 0.03 | 0.04 | 0.509 |
| Trend (pre) |  | < 0.001(-) | < 0.001(-) | 0.855 |
| Mean (post) |  | 0.89 | 0.89 | 0.531 |

|  |  |  |  |
| --- | --- | --- | --- |
| SD (post) | 0.03 | 0.04 | 0.003 |
| Trend (post) | < 0.001(+) | < 0.001(-) | 0.091 |
| **Sleep Fragmentation** |  |  |  |
| Mean (pre) | 0.12 | 0.13 | 0.691 |
| SD (pre) | 0.04 | 0.05 | 0.457 |
| Trend (pre) | < 0.001(-) | < 0.001(+) | 0.350 |
| Mean (post) | 0.13 | 0.12 | 0.546 |
| SD (post) | 0.04 | 0.05 | 0.017 |
| Trend (post) | < 0.001(-) | < 0.001(+) | 0.532 |
| **Sleep Stages** |  |  |  |
| REM Mean (pre) | 0.20 | 0.19 | 0.056 |
| REM SD (pre) | 0.06 | 0.07 | <0.001 |
| REM Trend (pre) | < 0.001(-) | < 0.001(-) | 0.321 |
| REM Mean (post) | 0.20 | 0.20 | 0.446 |
| REM SD (post) | 0.06 | 0.07 | 0.017 |
| REM Trend (post) | < 0.001(-) | < 0.001(-) | 0.094 |
| Light Mean (pre) | 0.65 | 0.65 | 0.643 |
| Light SD (pre) | 0.08 | 0.09 | <0.001 |
| Light Trend (pre) | < 0.001(-) | < 0.001(+) | 0.112 |
| Light Mean (post) | 0.65 | 0.65 | 0.535 |
| Light SD (post) | 0.08 | 0.09 | 0.175 |
| Light Trend (post) | < 0.001(-) | < 0.001(+) | 0.068 |
| Deep Mean (pre) | 0.15 | 0.15 | 0.080 |
| Deep SD (pre) | 0.05 | 0.05 | 0.114 |
| Deep Trend (pre) | < 0.001(+) | < 0.001(-) | 0.158 |
| Deep Mean (post) | 0.14 | 0.15 | 0.027 |
| Deep SD (post) | 0.05 | 0.05 | 0.061 |
| Deep Trend (post) | < 0.001(+) | < 0.001(-) | 0.581 |

Sleep stage features are based on participants with stage-mode devices (n = 244). Continuous variables are summarized as mean (SD = Standard Deviation), and “trend” reflects the linear slope over time. For sleep efficiency and fragmentation, slopes extremely close to zero are reported as “<0.001,” with a positive (+) or negative (-) sign to indicate the direction of change. Continuous variables were compared pain groups using Welch’s two-sample t-tests. P-values are reported to three significant digits; continuous summaries are shown to two decimals.

**Supplemental Table 3.** Comparison of model performance across demographic and multivariate models.

| Model | Train AUC | Test AUC | AIC | Sensitivity | Specificity |
| --- | --- | --- | --- | --- | --- |
| Demographics (Base model) | 0.636 | 0.584 | 243.53 | 0.032 | 1.00 |
| Physical Activity (Step counts) | 0.695 | 0.677 | 248.73 | 0.10 | 0.95 |
| Heart Rate | 0.724 | 0.596 | 239.65 | 0.23 | 0.90 |
| Sleep Features | 0.798 | 0.722 | 241.15 | 0.42 | 0.92 |
| Sleep Stage Features | 0.799 | 0.652 | 207.59 | 0.19 | 0.87 |
| Multivariate (All) | 0.753 | 0.679 | 239.92 | 0.26 | 0.92 |
| Multivariate (All - stepwise) | 0.720 | 0.648 | 227.07 | 0.26 | 0.92 |
| Multivariate (Stage-only) | 0.816 | 0.671 | 201.83 | 0.44 | 0.87 |
| Multivariate (Stage-only - stepwise) | 0.781 | 0.649 | 178.75 | 0.37 | 0.87 |

Multivariate (All) included the full cohort (n = 302), while Multivariate (Stage-only) only included participants with sleep stage-mode data available (n = 244).

